# Cumulative incidence of SARS-CoV-2 infection in the general population of the Valencian Community (Spain) after the surge of the Omicron BA.1 variant

**DOI:** 10.1101/2022.07.19.22277747

**Authors:** Jorge Camacho, Estela Giménez, Eliseo Albert, Joao Zulaica, Beatriz Álvarez-Rodríguez, Ignacio Torres, Luciana Rusu, Javier S. Burgos, Salvador Peiró, Hermelinda Vanaclocha, Ramón Limón, María Jesús Alcaraz, José Sánchez-Payá, Javier Díez-Domingo, Iñaki Comas, Fernando Gonzáles-Candelas, Ron Geller, David Navarro, the Valencian Vaccine Research Program (ProVaVac) study group

**Affiliations:** Microbiology Service, Clinic University Hospital, INCLIVA Health Research Institute, Valencia, Spain; Institute for Integrative Systems Biology (I2SysBio), Universitat de Valencia-CSIC, 46980, Valencia, Spain; General Directorate of Research and Healthcare Supervision, Department of Health, Valencia Government, Valencia, Spain; Foundation for the promotion of health and biomedical research of the Valencian Community (FISABIO), Valencia, Spain; General Directorate of Public Health, Department of Health, Valencia Government, Valencia, Spain; General Directorate of Healthcare. Department of Health, Valencian Government, Valencia, Spain; Preventive Medicine Service, Alicante General and University Hospital, Alicante, Spain; Alicante Institute of Health and Biomedical Research (ISABIAL), Alicante, Spain; Biomedicine Institute of Valencia, Spanish Research Council (CSIC); CIBER in Epidemiology and Public Health, Spain; Joint Research Unit “Infection and Public Health” FISABIO-University of Valencia, Valencia, Spain; Department of Microbiology, School of Medicine, University of Valencia, Valencia, Spain; Vice-President Foundation Research Institute in Public Services, Valencia, Spain; The Prince Felipe Research Center-CIPF-, Valencia, Spain; INCLIVA Health Research Institute, Valencia, Spain; Foundation for the promotion of health and biomedical research of the Valencian Community-FISABIO-, Valencia, Spain; CIBER in Epidemiology and Public Health, Spain; Joint Research Unit “Infection and Public Health” FISABIO-University of Valencia, Valencia, Spain; Institute for Integrative Systems Biology (I2SysBio), CSIC-University of Valencia, Valencia, Spain; Fundación Hospital Provincial de Castelló; Instituto de Investigación Sanitaria La Fe; Biotechnology Department, University of Alicante, Spain; Fundación Hospital General Universitario de València

**Keywords:** SARS-CoV-2, Seroprevalence, Cumulative incidence of SARS-CoV-2 infection, COVID-19 vaccine, natural infection, neutralizing antibodies, T cells

## Abstract

**Background:** Studies investigating the cumulative incidence of and immune status against SARS-CoV-2 infection provide valuable information for shaping public health decision-making.

**Methods:** The current cross-sectional, population-based study, conducted in April 2022 in the Valencian Community (VC), recruited 935 participants of all ages. Anti-SARS-CoV-2-Receptor Binding Domain-RBD-total antibodies and anti-Nucleocapsid (N)- IgGs were measured by electrochemiluminescence assays. To account for past SARS-CoV-2 infection the VC microbiology registry (RedMiVa) was interrogated. |Quantitation of neutralizing antibodies (NtAb) against the ancestral and Omicron BA.1 and BA.2 (sub)variants by an S-pseudotyped neutralization assay and for enumeration of SARS-CoV-2-S specific-IFNγ-producing CD4^+^ and CD8^+^ T cells by Intracellular Cytokine Staining assay was performed in a subset of participants (n=100 and 137, respectively).

**Findings:** The weighted cumulative incidence was 51□9% (95% CI, 48□7–55□1), and was inversely related to age. Anti-RBD total antibodies were detected in 906/931 (97□3%) participants, those vaccinated and SARS-CoV-2-experienced (VAC-ex;=442) displaying higher levels (*P*<0.001) than vaccinated/naïve (VAC-n;(n=472) and non-vaccinated/experienced (UNVAC-ex; n(n=63). Antibody levels correlated inversely with the time elapsed since receipt of last vaccine dose in VAC-n (Rho, -0□52; 95% CI, -0□59 to -0□45; *P*<0.001) but not in VAC-ex. NtAbs against Omicron BA.1 were detected in 94%, 75% and 50% of VAC-ex, VAC-n and UNVAC-ex groups, respectively, while in 97%, 84% and 40%, against Omicron BA.2. SARS-CoV-2-S-reactive IFN-γ T cells were detected in 73%, 75%, and 64% for VAC-ex, VAC-n, UNVAC-ex, respectively.

**Interpretation:** By April 2022 around half of the VC population had been infected with SARS-CoV-2 and due to extensive vaccination display hybrid immunity. The large percentage of participants with detectable functional antibody and T-cell responses against SARS-CoV-2, which may be cross-reactive to some extent, points towards lower expected severity than in previous waves.

**Funding:** This research was supported in part by the European Commission NextGenerationEU fund (CSIC’s Global Health Platform).

## INTRODUCTION

Spain has been severely affected by the SARS-CoV-2 pandemic, with more than 12.4 million microbiologically confirmed cases and almost 107,000 allegedly related deaths as of June 8^th^, 2022.^1^ The Valencian Community (VC), located along the Mediterranean coast in the east of the Iberian Peninsula, is the fourth most populous Spanish autonomous community with more than five million inhabitants. VC has reported around 1□5 million SARS-CoV-2 infection cases and almost 10,000 attributed deaths to date.^1^ Importantly, a large percentage of the VC population has received either a primary full vaccination series (93□9%) or a booster vaccination schedule (57□3%; about 95% in the over 60s).^2^ Due to the particular nature of SARS-CoV-2 infection with a large proportion of asymptomatic cases, limited testing among individuals with suspected COVID-19, and suboptimal contact tracing, registries of confirmed COVID-19 cases tend to underestimate the cumulative incidence of SARS-CoV-2 infection, as previously shown in the ENE-COVID study, a nationwide, population-based, seroepidemiological study conducted in Spain during 2020.^3^

A few serosurvey studies have been reported involving the general population and extending into the Omicron wave;^4-6^ these studies provide valuable information on the extent of transmission in the past and contribute towards understanding the future course of the pandemic and to shaping public health decision making. Here, as part of the ProVaVac Valencian COVID-19 vaccine research program launched by the VC government in March 2021, we conducted a population-based study aimed primarily at estimating the cumulative incidence of SARS-CoV-2 infection in the general VC population after the surge and spread of the Omicron BA.1 variant. Given that both SARS-CoV-2-Spike (S)-binding functional antibodies and S-reactive T cells elicited following vaccination or natural infection seemingly contribute to protection against severe COVID-19,^7-9^ we assessed as a secondary goal of the study the impact of COVID-19 vaccination, SARS-CoV-2 infection and time elapsed since these two events on anti-SARS-CoV-2-S(Spike)-Receptor Binding domain (RBD) total antibody levels in all participants. In this context, we also measured SARS-CoV-2-S Wuhan-Hu-1 (G614), Omicron BA.1 and BA.2 neutralizing Ab titers (NtAb) and SARS-CoV-2-S-reactive IFN-γ-producing CD4^+^ and CD8^+^ T-cell levels in a subsample of randomly selected subjects representative of the full recruited population.

## METHODS

### Study design and setting

The current cross-sectional, region-wide, population-based study was conducted in the primary care zones (PCZ) of the Valencia Health System) in April 2022, after the Omicron BA.1 variant had emerged and spread extensively within the VC. The sample size was calculated to be 998 participants for a precision of ±1% (between ±3 and ±5% in analyses by age and sex strata), assuming a prevalence of anti-RBD antibodies in the general population of 95% and a confidence level of 90%. The sample was increased by 5% (up to 1,043 blood samples) to compensate for possible losses, and was age- and sex-stratified proportionally to the distribution of the VC population, except for oversampling of ≥80 age group and reduction of the 35–49 and 50–64 age groups. For operational reasons, 100 PCZ were randomly selected to simplify sample transport logistics. Each PCZ was given a specific date on which to collect between 8 and 17 samples depending on the size of the population served, stratified by age/sex. Samples were obtained from an additional sodium heparin 5 cc tube collected from patients having blood drawn on doctor’s orders for any reason and meeting the age/sex selection criteria assigned to each center.

### Ethical statement

Because the survey was carried out under the epidemiological surveillance competences of the Valencian Community Autonomous Government (Law 10/2014, Dec 29 on Public Health of the Valencian Community, Spain), the Research Ethics Committee of the Foundation for the Health and Biomedical Research Promotion of the Valencian Community, Valencia, Spain (Research Ethics Committee of Public Health) waived the need for ethical approval or informed consent (Research Ethics Committee reference: 20220408/02). All personal data were processed under the standards of the Valencia Goverment Health Department, in accordance with the European data protection regulations and the Spanish laws on data protection

### Variables and definitions

Registered past SARS-CoV-2 infection was established by consulting the VC microbiology registry (RedMiVa) and taking into account the results of SARS-CoV-2 antibody assays performed, as described below. The vaccination status of participants was obtained from the VC vaccination registry. According to vaccination and SARS-CoV-2 infection status at the time of recruitment, participants were grouped into the following categories: vaccinated/experienced (VAC-ex, vaccinated individuals who had a record of a positive AIDT-active infection diagnostic test-result and/or serological evidence of past infection), vaccinated/naïve (VAC-n, vaccinated participants with no record or serological evidence of previous SARS-CoV-2 infection), unvaccinated/experienced (UNVAC-ex, unvaccinated population with history or serological evidence of prior SARS-CoV-2 infection), and unvaccinated/naïve (UNVAC-n, unvaccinated population with no history or serological evidence of prior SARS-CoV-2 infection). AIDTs included rapid antigen-detection assays targeting the N protein or commercially-available RT-PCRs.

### Immunological testing

SARS-CoV-2-RBD-total antibodies and nucleocapsid (N)-reactive IgG antibodies were measured in sera by the Roche Elecsys® Anti-SARS-CoV-2 S and Elecsys® Anti-SARS-CoV-2 N assays (Roche Diagnostics, Pleasanton, CA, USA), respectively, following the manufacturer’s recommendations. Values ≥0□4 BAU/ml and ≥1□0 cut-off index (COI), respectively were considered positive results. Sera returning BAU/ml values above the limit of quantitation were diluted appropriately and re-run. Neutralizing antibodies (NtAb) targeting the S protein were measured using a GFP-expressing vesicular stomatitis virus pseudotyped with the Wuhan-Hu-1, and Omicron BA.1 and BA.2 variants, as previously described,^10^ (Appendix). Sera resulting in less than 50% neutralization at the lowest dilution tested were ascribed a reciprocal neutralization titer of 10, equaling the limit of detection of the assay. SARS-CoV-2-S specific-IFNγ-producing CD4^+^ and CD8^+^ T cells were enumerated by whole-blood flow cytometry for intracellular cytokine staining [ICS] (BD Fastimmune, Becton Dickinson and Company Biosciences, San Jose, CA), as previously reported,^10,11^ (Appendix). Any frequency value after background subtraction was considered a positive (detectable) result and used for analysis purposes.

### Statistical analysis

Frequency comparisons for categorical variables were carried out using the Fisher exact test. Differences between medians were compared using the Mann–Whitney U-test, the Wilcoxon test, or Kruskal-Wallis test as appropriate. Two-sided exact *P*-values were reported. A *P*-value <0□05 was considered statistically significant. The total results are shown for the sample and weighted by the proportion of VC population in each age/sex stratum. The analyses were performed using SPSS version 20□0 (SPSS, Chicago, IL, USA) and STATA 17□0 (StataCorp, College Station, Texas, USA).

## RESULTS

### Participant characteristics

Out of 1,043 eligible participants, a total of 935 (89□6%), 524 (56%) female and 411 (44%) male subjects aged between 0–87 years (median, 51 years) were finally enrolled in the study (Supplementary Table 1). Nonattendance of participants (mostly children) matching predetermined requirements regarding sex and age prevented full recruitment, as planned. The number of participants from each VC province was balanced to the respective population size. Detailed information regarding the vaccine platforms used and the number of doses received by participants are shown in Supplementary Table 2. In summary, 812 (86□8%) participants had received a complete vaccine schedule (in most cases, Comirnaty® or Spikevax®), 39 (4□1%) were incompletely vaccinated and 84 (8□9%) were unvaccinated (48% children aged under 9 years) at the time of recruitment. The percentage of fully vaccinated participants increased with age. Booster vaccine doses were administered to 582 (69%) participants (homologous in 276 [47%, mostly children and elderly] and heterologous in the remaining 306 [53%]) (Table 1). The median time elapsed between receipt of the last vaccine dose and recruitment was 118 days (range, 2–428).

**Table 1.** COVID-19 Vaccination status among participants according to their age.

|  |  |  |  |  |
| --- | --- | --- | --- | --- |
| Age groups (no. of participants) | Vaccination status |  |  |  |
|  | no. of participants (%) | no. of participants (%) | no. of participants that received booster COVID-19 vaccine dose/s (%) | Non-vaccinated |

|  | that received an incomplete vaccine schedule | that received a complete vaccine schedule | All combined | Heterologous | Homologous | participants (%) |
| --- | --- | --- | --- | --- | --- | --- |
| All ages (935) | 39 (4) | 812 (89) | 582 (69) | 306 (52 □ 5) | 276 (47 □ 5) | 84 (9) |
| 0-9 (40) | 13 (33) | 8 (20) | 0 | - | - | 19 (48) |
| 10-19 (88) | 10 (11) | 68 (77) | 9 (12) | 7 (78) | 2 (22) | 10 (11) |
| 20-34 (150) | 10 (7) | 126 (84) | 63 (46) | 50 (79) | 13 (21) | 14 (9) |
| 35-49 (181) | 4 (2) | 158 (87) | 103 (64) | 79 (77) | 24 (23) | 19 (10) |
| 50-64 (174) | 1 (1) | 169 (97) | 135 (79) | 114 (84) | 21 (16) | 4 (2) |
| 65-79 (160) | 1 (1) | 152 (95) | 145 (95) | 48 (33) | 97 (67) | 7 (4) |
| >80 (142) | 0 (0) | 131 (92) | 127 (97) | 8 (6) | 119 (94) | 11 (8) |

**Table 2.**
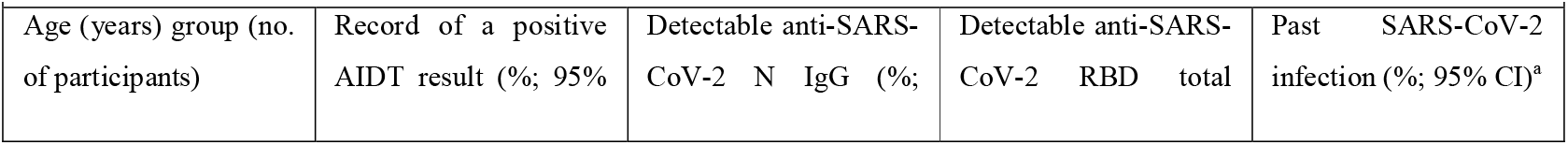
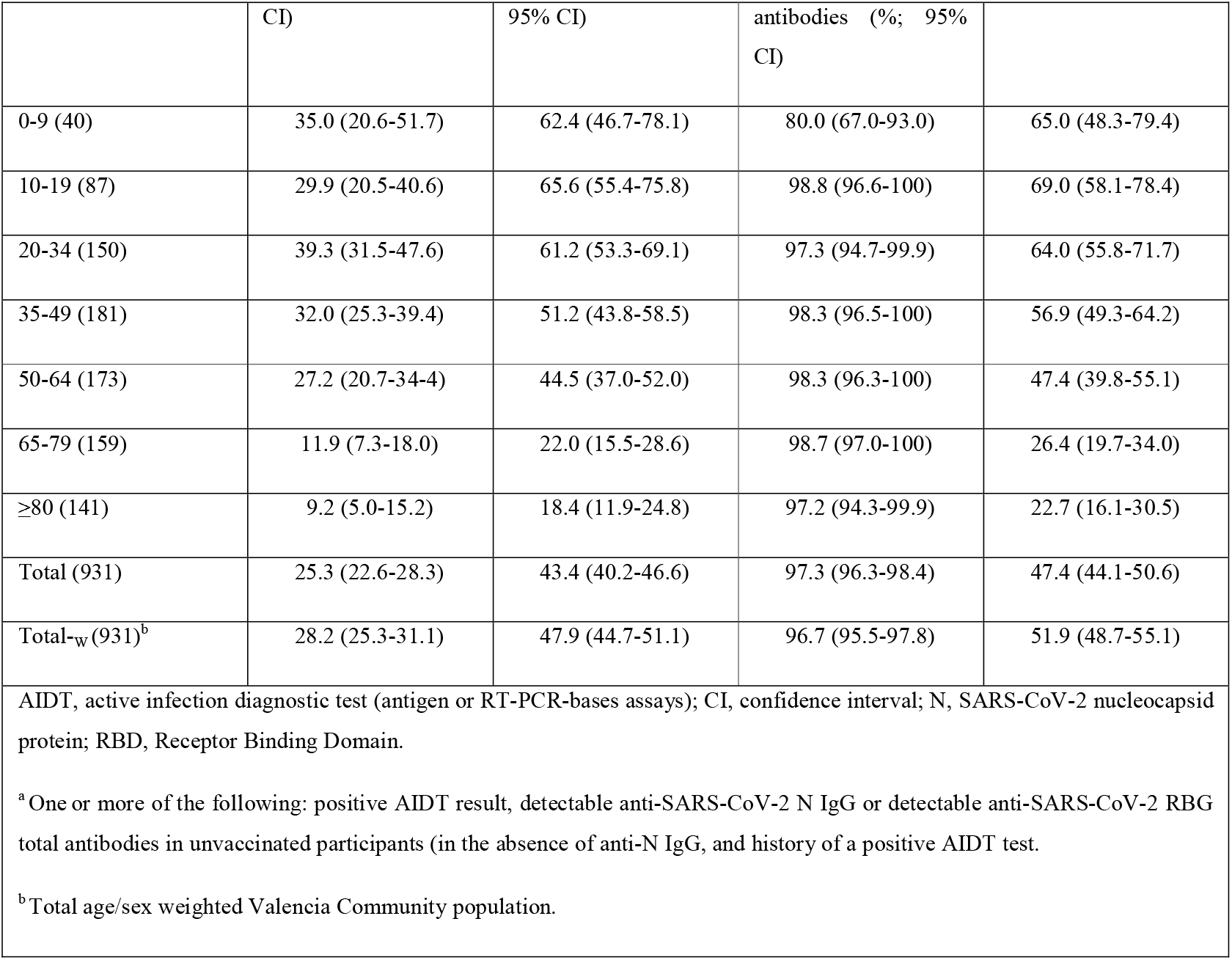
Cumulative incidence of past SARS-CoV-2 infection, as determined by active infection diagnostic test and/or serological assays by participants’ age.

|  |  |  |  |  |
| --- | --- | --- | --- | --- |
| Age (years) group (no. of participants) | Record of a positive AIDT result (%; 95% | Detectable anti-SARS-CoV-2 N IgG (%; | Detectable anti-SARS-CoV-2 RBD total | Past SARS-CoV-2 infection (%; 95% CI)<sup>a</sup> |

|  | CI) | 95% CI) | antibodies (%) 95% CI) |  |
| --- | --- | --- | --- | --- |
| 0-9 (40) | 35.0 (20.6-51.7) | 62.4 (46.7-78.1) | 80.0 (67.0-93.0) | 65.0 (48.3-79.4) |
| 10-19 (87) | 29.9 (20.5-40.6) | 65.6 (55.4-75.8) | 98.8 (96.6-100) | 69.0 (58.1-78.4) |
| 20-34 (150) | 39.3 (31.5-47.6) | 61.2 (53.3-69.1) | 97.3 (94.7-99.9) | 64.0 (55.8-71.7) |
| 35-49 (181) | 32.0 (25.3-39.4) | 51.2 (43.8-58.5) | 98.3 (96.5-100) | 56.9 (49.3-64.2) |
| 50-64 (173) | 27.2 (20.7-34.4) | 44.5 (37.0-52.0) | 98.3 (96.3-100) | 47.4 (39.8-55.1) |
| 65-79 (159) | 11.9 (7.3-18.0) | 22.0 (15.5-28.6) | 98.7 (97.0-100) | 26.4 (19.7-34.0) |
| ≥80 (141) | 9.2 (5.0-15.2) | 18.4 (11.9-24.8) | 97.2 (94.3-99.9) | 22.7 (16.1-30.5) |
| Total (931) | 25.3 (22.6-28.3) | 43.4 (40.2-46.6) | 97.3 (96.3-98.4) | 47.4 (44.1-50.6) |
| Total-w (931)<sup>b</sup> | 28.2 (25.3-31.1) | 47.9 (44.7-51.1) | 96.7 (95.5-97.8) | 51.9 (48.7-55.1) |
| AIDT, active infection diagnostic test (antigen or RT-PCR-bases assays); CI, confidence interval; N, SARS-CoV-2 nucleocapsid protein; RBD, Receptor Binding Domain.<br><sup>a</sup> One or more of the following: positive AIDT result, detectable anti-SARS-CoV-2 N IgG or detectable anti-SARS-CoV-2 RBG total antibodies in unvaccinated participants (in the absence of anti-N IgG, and history of a positive AIDT test.<br><sup>b</sup> Total age/sex weighted Valencia Community population. |  |  |  |  |

### Cumulative incidence of SARS-CoV-2 infection

Blood volume was insufficient for analyses in four participants, who were accordingly excluded, leaving 931 individuals included in the analyses detailed below.

Past SARS-CoV-2 infection was documented in a total of 442 (47□4%) participants (Table 2 and Supplementary Table 3), on the basis of either the presence of anti-SARS-CoV-2-N IgGs (n=404), a recorded positive AIDT result (n=237), or isolated detection of anti-SARS-CoV-2-RBD total antibodies inunvaccinated/SARS-CoV-2-naïve individuals (n=16). The crude cumulative incidence of prior SARS-CoV-2 infection in the VC was thus 47□4% (95% CI, 44□1–50□6), with no differences across the three VC provinces, whereas age/sex weighted cumulative incidence was 51□9% (95% CI, 48□7–55□1). The rate of past SARS-CoV-2 infection was inversely related to age; evidence of prior SARS-CoV-2 infection was found in more than two-thirds of subjects younger than 34 years old, in contrast to around one-fourth of those over 65 years old. 0Importantly, among participants with a documented positive AIDT result (n=237), 11% were diagnosed with SARS-CoV-2 infection in 2020 (n=25), 22% in the first semester of 2021 (n=52), 17% in the second semester of 2021 (n=40) and 51% in the first trimester of 2022 (n=120), time periods when the ancestral Wuhan-Hu-1 (including G614), Alpha, Delta and Omicron BA.1 variants, respectively, were dominant in the VC. Overall, the median time elapsed between microbiological diagnosis of SARS-CoV-2 infection and recruitment was 111 days (range, 8–591). For analysis purposes, participants were grouped as VAC-ex (n=379), VAC-n (n=472), UNVAC-ex (n=63) and UNVAC-n (n=21).

### Anti-SARS-CoV-2-RBD total antibodies

Overall, 906/931 (97□3%) had detectable anti-RBD total antibodies. Most individuals (23/25) with undetectable levels were UNVAC-n children. As shown in Figure 1 (A), median anti-RBD total antibody levels were significantly higher (*P*<0□001) in VAC-ex than in VAC-n participants (median, 23,120 BAU/ml; IQR, 10,186–46,196 vs. median, 9,023 BAU/ml; IQR, 3,844–20,329); this difference was noticed across most age ranges (Supplementary Figure 1). In turn, VAC-ex and VAC-n individuals displayed significantly higher (*P*<0.001) antibody levels than UNVAC-ex participants (median, 622 BAU/ml; IQR, 82–6,913).

**Figure 1.**
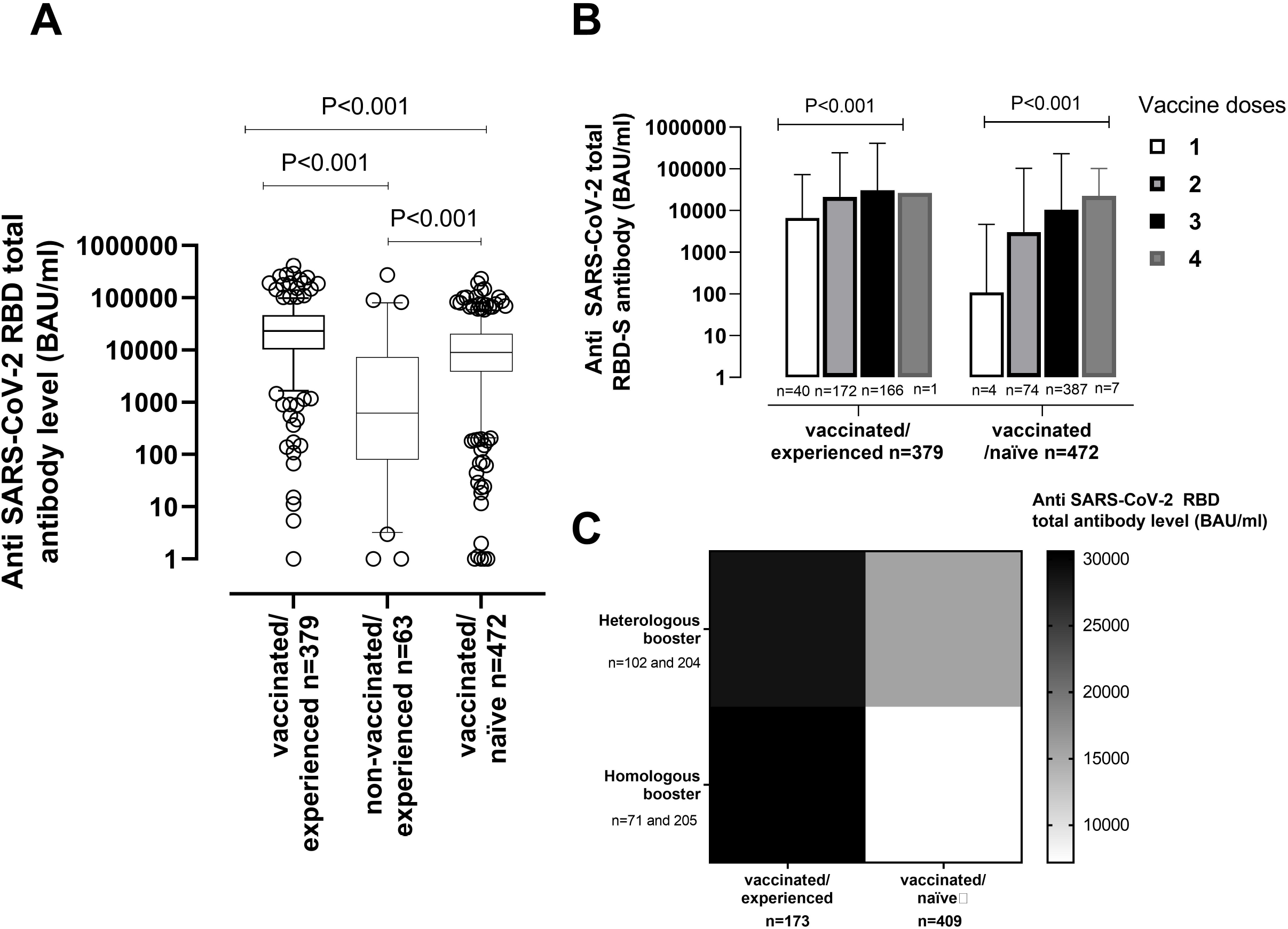
SARS-CoV-2-Receptor Binding Domain (RBD) total antibodies (in BAU/ml) in participants. (A) Box Whisker plots depicting serum antibody levels according to vaccination and SARS-CoV-2 infection status at time of recruitment. (B) Box Whisker plots depicting serum antibody levels in vaccinated/SARS-CoV-2-experienced and vaccinated/SARS-CoV-2-naïve participants by number of vaccine doses received. (C) Serum antibody levels in vaccinated/SARS-CoV-2-experienced and vaccinated/ SARS-CoV-2-naïve receiving one or more booster doses by booster type (homologous vs. heterologous). *P* values for statistical comparisons are shown when appropriate.

Compared to the VAC-n subgroup, individuals in the VAC-ex subgroup were younger (median 410 years; range 5–94 years; vs. 62 years; range, 5–95; *P*<0□001), and more frequently men (49% vs. 40%; *P*<0□02). Importantly, the time elapsed from the last vaccine dose to testing was shorter in VAC-n subjects than VAC-ex subjects (median 116 days; range, 0–428 vs. 150 days; range, 0–343; *P*<0.001), as the former subjects were more likely (*P*<0.001) to have received a booster vaccine dose than the latter (83□5 vs. 44□3%). In turn, VAC-ex and UNVAC-ex participants were matched for sex (*P*=0□62) and time since diagnosis of SARS-CoV-2 infection (median 92 days; range, 8–718; vs. median 82 days; range, 51–516; *P*=0□25). Nevertheless, the UNVAC-ex subgroup comprised more participants under 9 years than the VAC-ex group, while the VAC-ex group included more subjects aged 50–64 years (*P*<0□001).

Overall, the number of vaccine doses received had a direct impact on level of anti-RBD total antibodies, irrespective of SARS-CoV-2 infection status (Figure 1B). However, no correlation was found between the time elapsed since receipt of the last vaccine dose and anti-RBD total antibody levels (rho -0□02; 95% CI, -0□13 to 0□07; *P*=0□57) in VAC-ex individuals, whereas a moderate inverse correlation (Rho, -0□52; 95% CI, - 0□59 to -0□45; *P*<0□001) was observed in VAC-n (Figure 2). Interestingly, in VAC-ex participants with a recorded positive AIDT result [n=220] (Figure 3), the time elapsed since documentation of SARS-CoV-2 infection correlated weakly (inversely) with anti-RBD antibody levels (Rho, -0□30; 95% CI -□42 to -0□17; *P*<0□001).

**Figure 2.**
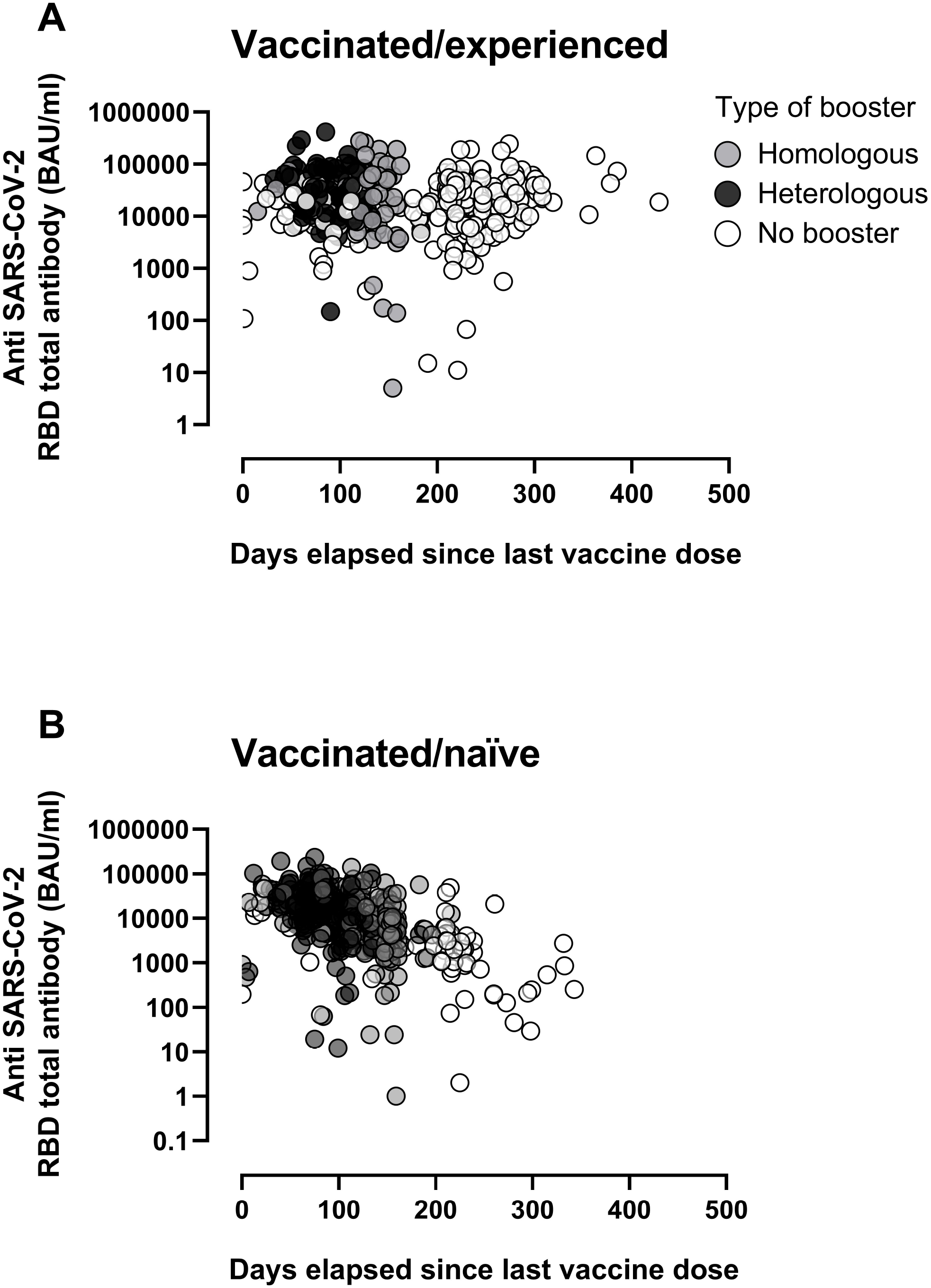
SARS-CoV-2-Receptor Binding Domain (RBD) total antibodies (in BAU/ml) in vaccinated/SARS-CoV-2-experienced (A) and vaccinated/SARS-CoV-2-naïve (B) participants according to the vaccination schedule (full vaccination with no booster dose, homologous booster dose and heterologous booster dose) and the time elapsed since receipt of the last vaccine dose.

**Figure 3.**
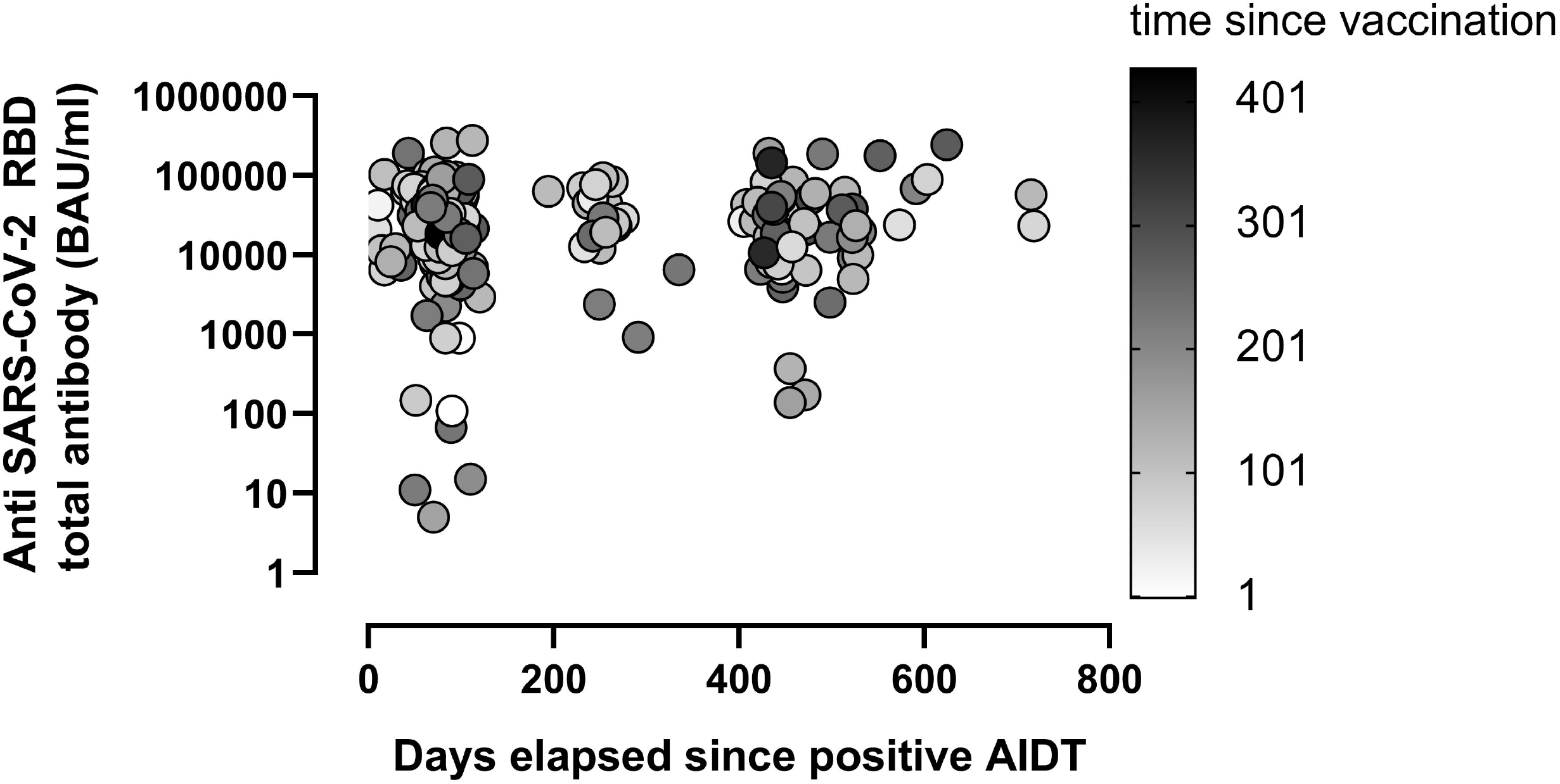
SARS-CoV-2-Receptor Binding Domain (RBD) total antibodies (in BAU/ml) in vaccinated/SARS-CoV-2 experienced participants with a history of a positive active infection diagnostic test (AIDT) according to the time elapsed since documentation of the positive result and time since receipt of the last vaccine dose.

Among those receiving a booster vaccine dose, heterologous booster shots resulted in increased anti-RBD antibody levels compared to homologous schedules in SARS-CoV-2-naïve, but not SARS-CoV-2-experienced participants, regardless of age and sex (Figure 1C).

Finally, subtle age-related differences in anti-RBD total antibody levels were noticed in both VAC-ex and VAC-n subgroups (Supplementary Figure 1), which were likely related to dissimilarities across age groups in the number of vaccine shots received and time elapsed since administration of the last vaccine dose.

### Neutralizing antibodies against the ancestral Wuhan-Hu-1 and Omicron subvariants

A total of 100 randomly selected participants across all age groups were screened for the presence of NtAb against SARS-CoV-2 (sub)variants. The main characteristics of the subjects evaluated are shown in Supplementary Table 4.

The proportion of participants displaying NtAbs against Wuhan-Hu-1 variant was 100%, 93% and 40% in the VAC-ex, VAC-n and UNVAC-ex groups, respectively; for Omicron BA.1 the percentages were 94%, 75% and 50%, and for Omicron BA.2, 97%, 84% and 40%, respectively.

Overall, NtAb titers were lower against Omicron BA-1 and BA.2 than Wuhan-Hu-1 across all comparison groups (Figure 4). In addition, NtAb targeting all (sub)variants screened were higher in VAC-ex than in VAC-n individuals, although statistical significance was reached for those against Wuhan-Hu-1 (*P*=0□05) and BA.1 (*P*<0□01), but not against BA.2 (*P*=0□74). Both VAC-ex and VAC-n displayed higher NtAb titers than UNVAC-ex participants (*P*=0□001 for Wuhan Hu-1; *P*=0□052 for BA.1, and *P*=0□003 for BA.2). Median levels of NtAb binding Omicron BA.1 and BA.2 were comparable in participants across all groups (Figure 4).

**Figure 4.**
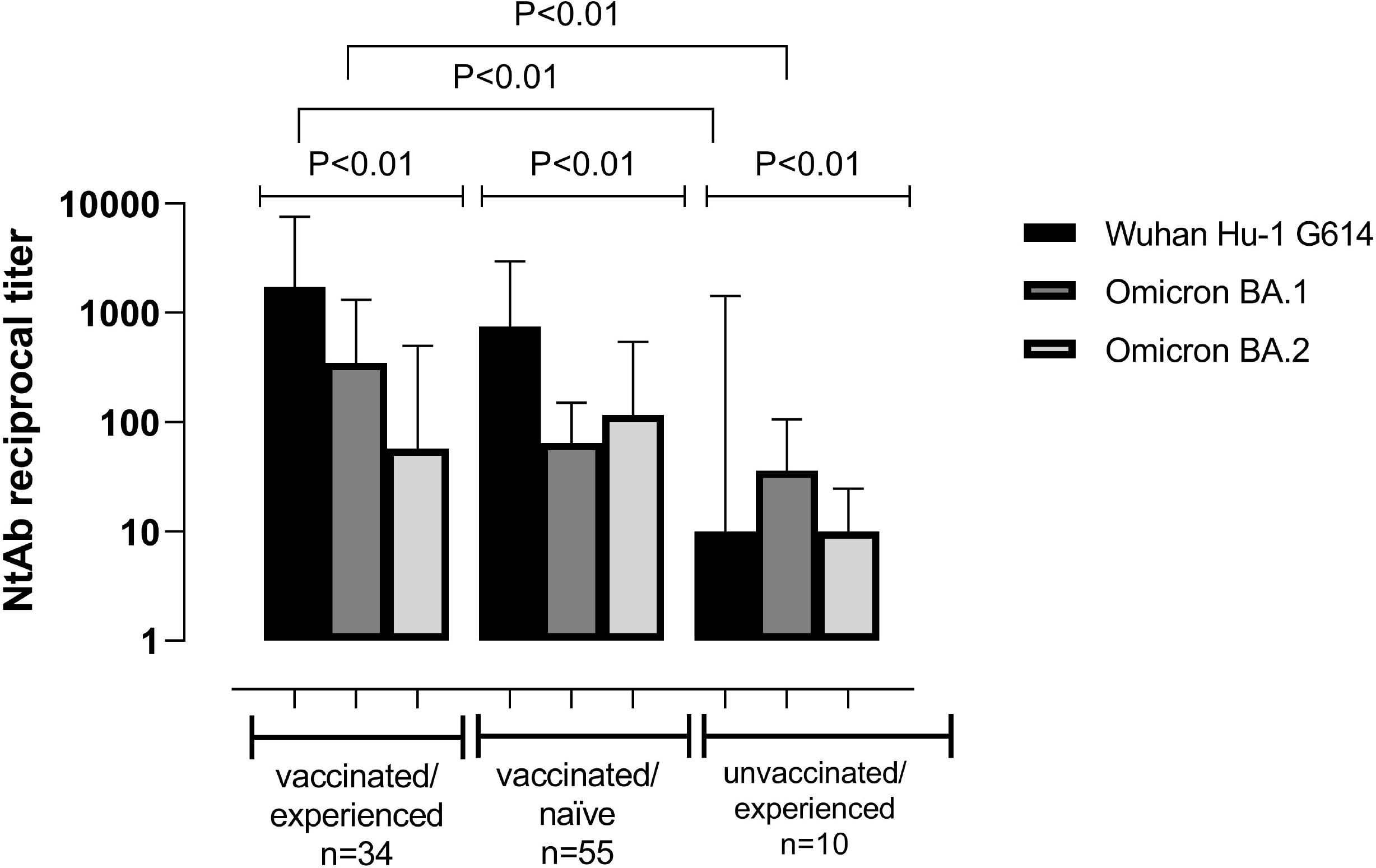
Serum neutralizing antibody titers (reciprocal IC_50_) against SARS-CoV-2 Wuhan-Hu-1, Omicron BA.1 and BA.2 (sub)variants in vaccinated/SARS-CoV-2 experienced, vaccinated/SARS-CoV-2 naïve participants and unvaccinated/SARS-CoV-2 experienced participants. Vaccinated/experienced, vaccinated/naïve and non-vaccinated/experienced participants were matched for sex and age, and vaccinated participants for booster type. Nevertheless, time since receipt of the last vaccine dose was significantly shorter (*P*<0.001) in vaccinated/naïve than in vaccinated/experienced participants (median 116 days; range, 29-298; vs. 162 days; range, 0-308). *P* values for statistical comparisons are shown.

Overall, comparable NtAb titers against all (sub)variants tested were seen across participants either boosted or not with an additional vaccine dose (Supplementary Table 5). Nevertheless, a significant although weak inverse correlation between NtAb titers against Wuhan-Hu-1 and Omicron BA.1 and time elapsed since the receipt of the last vaccine dose was observed in VAC-n but not VAC-ex individuals (Supplementary Table 6).

### SARS-CoV-2-Spike T-cell responses

We enumerated SARS-CoV-2-S-reactive IFN-γ CD4^+^ and CD8^+^ T cells in 137 (14□7%) participants (Supplementary Table 4). Overall, 101 (73□7%) participants had detectable SARS-CoV-2-S-IFN- γ T cells: either CD4^+^ (n=15; 14□9%), CD8^+^ (n=41; 40□6%), or both (n=45; 44□6%). In detail, the figures were 73%, 75%, and 64% for VAC-ex, VAC-n, UNVAC-ex, respectively. Median frequencies of both T-cell subsets were comparable across subgroups (*P*=0□92 for CD8^+^ T cells and *P*=0□80 for CD4^+^ T cells) (Figure 5)0. Likewise, neither age (*P*=0□86) nor sex (*P*=0□57) impacted on the median frequencies of the two T-cell subsets. In both SARS-CoV-2 T-cell subsets frequencies appeared comparable irrespective of time elapsed since the last vaccine dose (Supplementary Table 6) or whether booster vaccine doses had been received (Supplementary Table 5), in both VAC-ex and VAC-n individuals.

**Figure 5.**
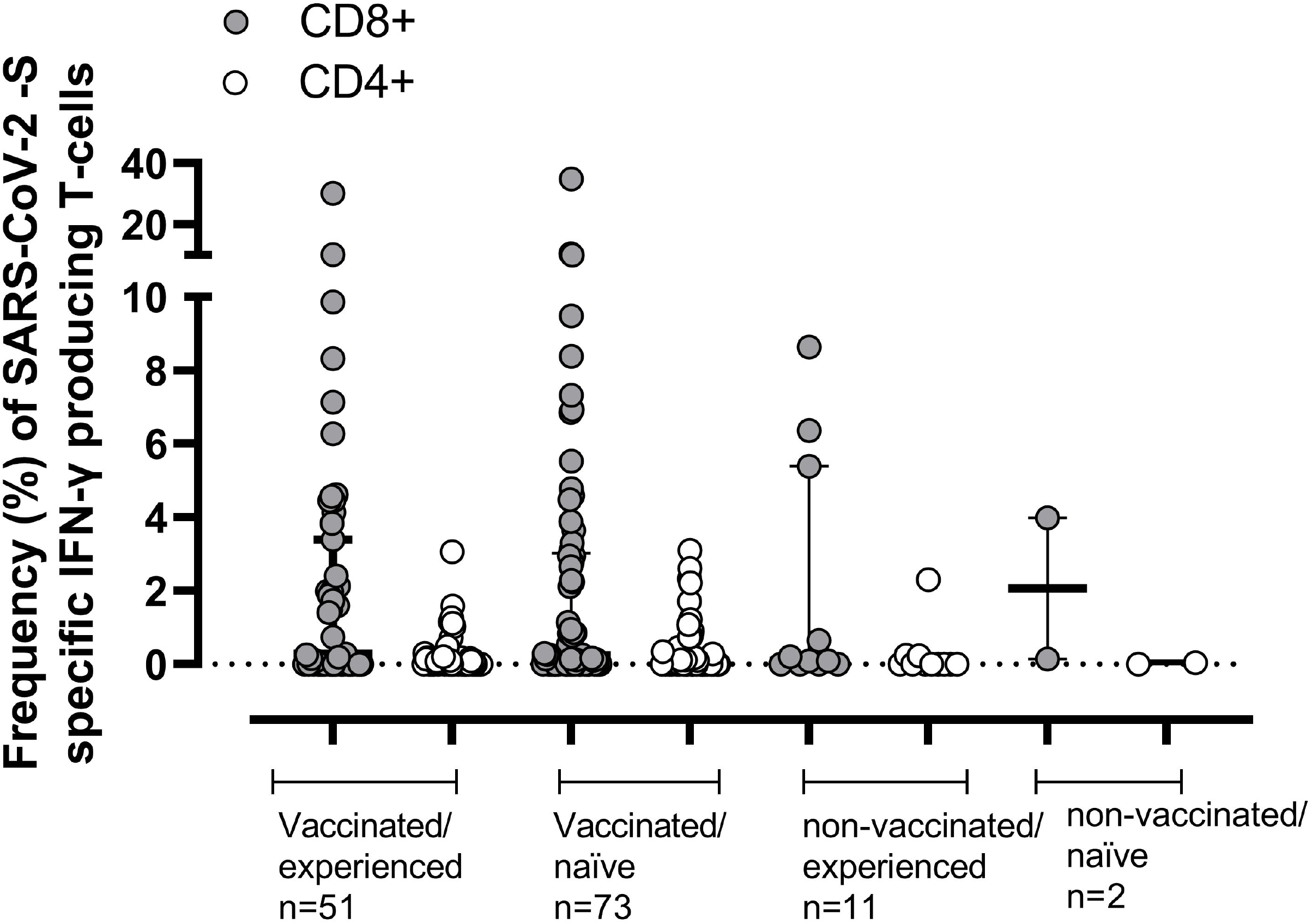
Frequency of SARS-CoV-2-Spike(S)-reactive IFN-γ-producing CD4^+^ and CD8^+^ T cells in vaccinated/SARS-CoV-2 experienced, vaccinated/SARS-CoV-2 naïve participants, unvaccinated/SARS-CoV-2 experienced and unvaccinated/SARS-CoV-2 naïve participants. Vaccinated/experienced, vaccinated/naïve and unvaccinated/experienced participants were balanced for sex and age, and for vaccinated individuals for booster type; nevertheless, time since receipt of the last vaccine dose was significantly lower (*P*<0.001) in vaccinated/naïve than in vaccinated/experienced participants (median 113 days; range, 0-298; vs. 175 days; range, 0-308).

## DISCUSSION

Knowledge of the current immune status against SARS-CoV-2 in the general population is central to predicting how the pandemic will evolve in the near future. In communities with extensive COVID-19 vaccine coverage, such as the VC (and Spain), determining the number of people who have contracted SARS-CoV-2 infection is of paramount relevance, since hybrid immunity (resulting from natural infection and vaccination) may outperform immunity from vaccination or (re)infection alone in terms of protection against SARS-CoV-2 infection and severe COVID-19.^12-14^

Only a handful of seroepidemiological studies have been performed to date in the general population extending into the Omicron wave.^4-6^ The proportion of infected individuals has dramatically increased following the surge of the Omicron sub-lineages, due to their increased transmissibility and escape from vaccine or infection-induced immunity.^15^ Moreover, as the percentage of Omicron-infected paucisymptomatic or asymptomatic individuals appears to be high, the number of underreported infections that are not captured by community surveillance systems may be significantly greater.

Here, we conducted a cross-sectional serosurvey to estimate the prevalence of SARS-CoV-2 antibodies in the general population of the VC after the surge in Omicron BA.1 variant cases and retrieved from the RedMiVa VC registry documented prior SARS-CoV-2 infection among the recruited population. Our results can likely be extrapolated to other autonomous communities in Spain.^3^ We estimated an age/sex weighed cumulative incidence of prior SARS-CoV-2 infection of 51□9% (95% CI, 48□7– 55□1). While this figure was comparable across males and females, age-related differences were noticed, the highest rate occurring in individuals under 34 years old and the lowest in those aged 65 or older. Such differences could be explained by dissimilarities in the rate of full or booster vaccination and social behavior across age groups. Interestingly, around 9% of participants lacked anti-SARS-CoV-2-N antibodies despite documentation of previous SARS-CoV-2 infection, suggesting, in line with previous findings,^16^ that assessment of anti-N antibodies alone may underestimate the incidence of SARS-CoV-2 infection.

Evaluation of SARS-CoV-2 immune status according to vaccine schedule, time elapsed since vaccination, and previous virus exposure also provides valuable information to model the course of the pandemic. Here, we measured serum anti-RBD total antibody levels as a surrogate for neutralizing activity of sera,^6,7^ in all participants, and NtAb titers and the frequency of peripheral blood SARS-CoV-2-S-reactive IFN-γ CD4^+^ and CD8^+^ T cells in a randomly selected subset representing the entire study population. The following facts are relevant for interpreting the data. First, the median time elapsed from receipt of last vaccine dose to testing was around 4 months; second, among participants with a documented positive AIDT result, the median time elapsed between SARS-CoV-2 diagnosis and recruitment was 111 days overall; third, as stated above approximately 50% of SARS-CoV-2 infections in recruited subjects with a positive AIDT result were seemingly caused by the Omicron BA.1 variant.

In this setting, 97% of participants had detectable anti-RBD total antibodies, whose levels were increased in VAC-ex individuals compared to VAC-n participants across virtually all age groups. This occurred irrespective of the time elapsed since diagnosis of SARS-CoV-2 infection, and despite the fact that the VAC-n group had received the last vaccine dose (in most cases a booster dose) more recently than the VAC-ex one. These data can be interpreted as suggesting that hybrid immunity provides stronger functional antibody responses than vaccination alone, as previously reported.^17-19^ Moreover, such responses may be more durable in the VAC-ex, as evidenced by an inverse correlation between the time elapsed since last vaccine dose and anti-RBD total antibody levels in VAC-n but not VAC-ex participants. VAC-ex individuals also displayed greater anti-RBD antibody levels than UNVAC-ex individuals matched by time of SARS-CoV-2 diagnosis, suggesting that antibody waning may occur at a faster rate in these latter individuals.

Our data on NtAb experiments seemed to support the assumption that hybrid immunity provides a more robust and perhaps more durable functional antibody response against Wuhan-Hu-1 and Omicron BA.1 than is elicited by either vaccination or natural infection alone. A key observation of our study was the differential effect of a booster dose on antibody levels across VAC-ex and VAC-n individuals. In effect, the data indicated that following the vaccine booster, both anti-RBD antibody levels and NtAb titers increased to a much lesser extent in VAC-ex than VAC-n, suggesting that in the general population VAC-n may benefit more than VAC-ex from receiving additional vaccine doses. This could have important implications in the design of vaccination policies in the near future. Regarding the impact of booster vaccine doses on humoral immunity, we showed that heterologous mRNA vaccine boosters elicited higher levels of anti-RBD total antibodies than homologous schedules, as previously reported.^20,21^ However, a similar analysis could not be performed for NtAb due to the small sample size.

Our finding that a large percentage of vaccinated participants had either prior SARS-CoV-2 infection or detectable NtAb against Omicron BA.1 (94% and 75% respectively) is compatible with the idea that both Wuhan-based vaccine platforms and breakthrough infections elicit cross-reactive antibodies against SARS-CoV-2 variants of concern^22,23^ and that Omicron BA.1 breakthrough infection may induce sublineage-specific NtAb in addition to recall of memory B cells established by prior vaccination.^24-26^

Currently, Omicron BA.2 has replaced BA.1 to become the dominant variant in the VC and Spain. In our study population, 50% of VAC-ex and 40% of VAC-n individuals displayed detectable NtAb responses against Omicron BA.2. Interestingly, median levels of NtAb binding Omicron BA.1 and BA.2 were comparable in participants across all study subgroups, suggesting a substantial degree of B-cell-epitope cross-reactivity across subvariants, as previously observed.^27^

Detectable SARS-CoV-2 T-cell responses (particularly CD4^+^ T cells are known to persist up to 8 months following natural infection or vaccination.^28,29^ Our data seem to support this notion, in that either SARS-CoV-2-S-IFN-γ CD4^+^, CD8^+^ T cells, or both could be detected in 75% of participants at a median of 4 months after receipt of the last vaccine dose. In effect, SARS-CoV-2-S IFN-γ T-cell responses may maintain relatively stable over time, since frequencies of both T-cell subsets were not correlated with the time elapsed since last vaccine dose. Moreover, administrating a booster dose had a minimal impact on the magnitude of T cell responses. Interestingly, in contrast to our observations in functional antibodies, we found no differences between VAC-ex and VAC-n in the frequencies of either T-cell subsets.

The current study has several limitations. First, bias may have been introduced by excluding some PCZs and by only including individuals scheduled for blood test, whichare likely sicker than the general population, both potentially translating into differential behaviors related to SARS-CoV-2 transmission. Second, participants were not asked about whether or not they had recently experienced COVID-19-related symptoms. Third, seroprevalence studies cannot account for reinfections and therefore may underestimate the cumulative number of SARS-CoV-2 infections. Fourth, sequencing profiling of SARS-CoV-2 variants was not performed. Fifth, an unstandardized flow cytometry ICS assay was used for enumerating SARS-CoV-2-S-reactive T cells. Moreover, T-cell functionalities other than IFN-γ production were not explored and Wuhan-Hu-1-based instead of VOCs-adapted overlapping peptide libraries were used.

In summary, our study indicates that by April 2022 around half of the VC population had been infected with SARS-CoV-2 and due to extensive vaccination exhibit hybrid immunity. The Valencian Community is currently experiencing a rise in the incidence of SARS-CoV-2 infection, mainly involving the BA.5 omicron sublineage. The large percentage of participants with detectable functional antibody and T-cell responses against SARS-CoV-2 which may be cross-reactive with this subvariant to some extent points towards lower expected severity than in previous waves. Although still to be confirmed in wider studies, this aspect is relevant for decision-making in public health.

## Data Availability

All data produced in the present study are available upon reasonable request to the corresponding author

## CONTRIBUTORS

JC, EG, EA, JZ, IT, LR, BAR, MJA, IC, FG-C and RG carried out antibody and T-cell assays and validated and analyzed the data. RL was responsible for the logistics of the study. JSB, HV, SP, JS-P, JD-D, and DN: conceptualization, study design and data interpretation. SP and DN wrote the manuscript. All authors reviewed the final version of the manuscript.

## ACKNOWLEDGMENTS

We are grateful to the staff and nurses of primary care centers participating in this study. We also thank Ana Berenguer, General Director of Analysis and Public Policies of the Presidency of the Generalitat. Eliseo Albert (Juan Rodés Contract; JR20/00011) Estela Giménez (Juan Rodés Contract, JR18/00053) and Ignacio Torres (Río Hortega Contract; CM20/00090) hold contracts funded by the Carlos III Health Institute (co-financed by the European Regional Development Fund, ERDF/FEDER). Ron Geller holds a Ramon y Cajal fellowship from the Spanish Ministry of Economics and Competitiveness (RYC-2015-17517).

## Data Sharing

The data that support the findings of this study are available from the corresponding author DN upon reasonable request.

## FIGURE LEGENDS

**Supplementary Figure 1**. SARS-CoV-2-Receptor Binding Domain (RBD) total antibodies (in BAU/ml) in vaccinated/SARS-CoV-2 experienced and vaccinated/SARS-CoV-2 naïve vaccinated participants, according to their age. *P* values for statistical comparisons are shown.

