## Supplementary material for "Cumulative incidence of SARS-CoV-2 infection in the general population of the Valencian Community (Spain) after the surge of the Omicron BA.1 variant"


- ✓ ***Study design and Setting…page 2***
- ✓ ***Immunological assays…page 2-4***
- ✓ ***Supplementary Table 1---page 5***
- ✓ ***Supplementary Table 2…page 6***
- ✓ ***Supplementary Table 3…page 7***
- ✓ ***Supplementary Table 4…page 8***
- ✓ ***Supplementary Table 5…page9***
- ✓ ***Supplementary Table 6…page10***
- ✓ ***Supplementary Figure 1…page 11***

### *Study design and Setting*

The study was conducted in the primary care zones (PCZ) of the Valencia Health System (VHS), an extensive public network of hospitals, primary care centers, and other facilities managed by the regional government, which provides universal free healthcare services (except for drug copayment) to 97% of the regions' populations. The VHS is organized in 24 healthcare departments served by a public general hospital, and 241 PCZ served by at least one primary healthcare center, most offering a blood draw service. The VHS Integrated Database (VID) is a comprehensive information system with a unique personal identifier for the whole registered population, including temporary residents and people without VHS coverage. The VID includes a microbiological registry (RedMiVa), with the results of all the microbiological tests carried out in all the laboratories of the VC public network. In the case of COVID-19 it also includes all tests carried out by private laboratories (but not the antigen-based tests carried out by patients at home) and a vaccination registry, which incorporates all vaccines administered in the region.

For operational reasons, we excluded 46 PCZs with less than 8,000 inhabitants, which constituted 4.0% of the total VC population, mainly remote areas or areas served by centers offering blood draw services only a few days a month. Of the remaining 195 PCZ, 100 were randomly selected to simplify sample transport logistics.

### *Immunological assays*

Anti-RBD total antibody and T-cell assays were run at the Microbiology Service of the Hospital Clínico Universitario of Valencia (HCU), whereas NtAb were measured at the Institute for Integrative Systems Biology (I2SysBio), Universitat de Valencia-CSIC.

Pseudotype vesicular stomatitis virus (VSV)-based neutralization assays were performed as previously described [12,13], with several modifications. First, to improve pseudotyping efficiency, C-terminal 19 amino acids were deleted by PCR in a plasmid encoding a codon-optimized Wuhan spike gene (pCG1-Wuhan-S). Similarly, a commercially available plasmid encoding a codon-optimized BA.1 spike gene (Genescript MC_0101274) was modified to remove the terminal 19 amino acids and the open reading frame transferred to the pCG1 plasmid using NEBuilder HiFi DNA assembly kit (NEB). For BA.2, a codon-optimized plasmid encoding the BA.2 spike gene lacking the terminal 18 amino acids (InvivoGen catalog code: plv-spike-v12) was PCR amplified and ligated to the pCG1 plasmid using NEBuilder HiFi DNA assembly kit (NEB). All inserts were verified by Sanger sequencing. The neutralization capacity of circulating antibodies (NtAb) against the different SARS-CoV-2 S variants was then assessed using this GFP-expressing VSV pseudotyped virus in A549-ACE2 TMPRSS2 cells (InvivoGen catalog code: a549-hace2tps). All tests were done in duplicate using 5-fold dilutions starting at a 1:20 dilution, with ~300-500 focus forming units per well. Following 16 hours of infection, the GFP signal in each well was quantified using a live-cell microscope (Incucyte SX5, Sartorius). Background fluorescence from uninfected wells was subtracted from all infected wells, and the GFP fluorescence in each antibody-treated dilution was standardized to the average fluorescence observed in mock-treated wells. Any value resulting in a relative GFP signal of <0.001 versus untreated virus was assigned a value of 0.001 to eliminate negative values. Finally, the reciprocal antibody dilution resulting in 50% virus neutralization was calculated using the drc package (version 3.0-1) in R via a three-parameter log-logistic regression model (LL.3 model). Sera resulting in less than 50% neutralization at the lowest dilution tested were arbitrarily ascribed a reciprocal antibody titer of 10.

For T-cell immunity analyses, heparinized whole blood (0.5 ml) was simultaneously stimulated for 6 h with two sets of 15-mer overlapping peptides (11-mer overlap) encompassing the SARS-CoV-2 Spike (S) glycoprotein (S1, 158 peptides and S2, 157 peptides) at a concentration of 1 μg/ml per peptide, in the presence of 1 μg/ml of costimulatory monoclonal antibodies (mAbs) to CD28 and CD49d. Peptide mixes were obtained from JPT peptide Technologies GmbH (Berlin, Germany). Samples mock-stimulated with phosphate-buffered saline (PBS)/dimethyl sulfoxide and costimulatory antibodies were run in parallel. Brefeldin A (10 μg/ml) was added for the last 4 h of incubation. Blood was then lysed (BD FACS lysing solution) and frozen at −80°C until tested. On the day of testing, stimulated blood was thawed at 37°C, washed, permeabilized (BD permeabilizing solution) and stained with a combination of labeled mAbs (anti-IFNγ-FITC, anti-CD4-APC-H7, anti-CD8-PerCP-Cy5.5, and anti-CD3-APC) for 1 h at room temperature (Supplementary Table 2). Appropriate positive (phytohemagglutinin) and isotype controls were used. Cells were then washed, resuspended in 200 μL of 1% paraformaldehyde in PBS, and analyzed within 2 h on an FACSCanto flow cytometer using DIVA v8 software (BD Biosciences Immunocytometry Systems, San Jose, CA). CD3+/CD8+ or CD3+/CD4+ events were gated and then analyzed for IFN-γ production. All data were corrected for background IFN-γ production (FITC-labelled isotype control antibody). The data are expressed as the number of SARS-CoV-2-reactive IFN-γ-producing CD4+ or CD8+ T cells relative to the absolute number of CD4+ and CD8+ T cells, respectively, x100 (%). Any frequency value of SARS-CoV-2-reactive IFN-γ-producing CD4+ or CD8+ T cells after background substraction was considered a positive (detectable) result and used for analysis purposes.

| Supplementary Table 1. Demographics of study participants | | | | |
|---|---|---|---|---|
| **Parameter** | **Valencian Community provinces** | | | |
| | **All (n=935)** | **Valencia (n=547)** | **Alicante (n=323)** | **Castellón (n=65)** |
| **Sex (%)** | | | | |
| Male | 411 (44) | 241 (44.1) | 145 (44.9) | 25 (38.5) |
| Female | 524 (56) | 306 (55.9) | 178 (55.1) | 40 (61.5) |
| **Age groups in years (%)** | | | | |
| 0-9 | 40 (4) | 22 (4) | 15 (4.6) | 3 (4.6) |
| 10-19 | 88 (9) | 57 (10.4) | 24 (7.4) | 7 (10.8) |
| 20-34 | 150 (16) | 89 (16.3) | 51 (15.8) | 10 (15.4) |
| 35-49 | 181 (19) | 106 (19.4) | 63 (19.5) | 12 (18.5) |
| 50-64 | 174 (19) | 99 (18.1) | 66 (20.4) | 9 (13.8) |
| 65-79 | 160 (17) | 88 (16.1) | 60 (18.6) | 12 (18.5) |
| >80 | 142 (15) | 86 (15.7) | 44 (13.6) | 12 (18.5) |

| **Supplementary Table 2. Vaccine platforms and number of doses administered to study participants** | | | | | |
|---|---|---|---|---|---|
| **Vaccine platform** | **Type of vaccine** | **Number of vaccine doses (%)** | | | |
| | | **At least one** | **Two** | **Three** | **Four** |
| Comirnaty® (BioNTech /Pfizer) | RNA | 630 (74) | 597 (74) | 220 (39) | 1 (12) |
| Spikevax® (Moderna / Lonza) | RNA | 106 (12) | 134 (17) | 341 (61) | 7 (88) |
| Vaxzevria® (Oxford / AstraZeneca) | Non-replicating viral vector | 86 (10) | 75 (9) | - | - |
| Jcovden® (JandJ / Janssen) | Non-replicating viral vector | 26 (3) | - | - | - |
| CoronaVac® (Sinovac Biotech) | Inactivated | 1 (0.1) | 1 (0.1) | - | - |
| Convidecia® (CanSinoBIO) | Recombinant viral vector | 1 (0.1) | - | - | - |
| Sputnik V® (Gamaleya) | Recombinant viral vector | 1 (0.1) | - | - | - |

**Supplementary Table 3. Cumulative incidence of past SARS-CoV-2 infection according to the age and vaccination status of participants**

| Age (years) group (no. of participants) | Vaccinated | | | | | | Non-vaccinated | | | | | |
|---|---|---|---|---|---|---|---|---|---|---|---|---|
| | **Past infection[a]** | | | **Naïve** | | | **Past infection[a]** | | | **Naïve** | | |
| | **%** | **95%CI** | | **%** | **95%CI** | | **%** | **95%CI** | | **%** | **95%CI** | |
| 0-9 (40) | 35.0 | 20.6 | 51.7 | 17.5 | 7.3 | 32.8 | 30.0 | 16.6 | 46.5 | 17.5 | 7.3 | 32.8 |
| 10-19 (87) | 58.6 | 47.6 | 69.1 | 29.9 | 20.5 | 40.6 | 10.3 | 4.8 | 18.7 | 1.1 | 0.0 | 6.2 |
| 20-34 (150) | 56.7 | 48.3 | 64.7 | 34.0 | 26.5 | 42.2 | 7.3 | 3.7 | 12.7 | 2.0 | 0.4 | 5.7 |
| 35-49 (181) | 48.1 | 40.6 | 55.6 | 41.4 | 34.2 | 49.0 | 8.8 | 5.1 | 13.9 | 1.7 | 0.3 | 4.8 |
| 50-64 (173) | 46.2 | 38.6 | 54.0 | 51.4 | 43.7 | 59.1 | 1.2 | 0.1 | 4.1 | 1.2 | 0.1 | 4.1 |
| 65-79 (159) | 23.3 | 16.9 | 30.6 | 72.3 | 64.7 | 79.1 | 3.1 | 0.1 | 7.2 | 1.3 | 0.1 | 4.5 |
| ≥80 (141) | 17.0 | 11.2 | 24.2 | 75.2 | 67.2 | 82.1 | 5.7 | 2.5 | 10.9 | 2.1 | 0.4 | 6.1 |
| Total (931) | 40.6 | 37.4 | 43.8 | 50.4 | 47.1 | 53.6 | 6.7 | 5.2 | 8.6 | 2.2 | 1.4 | 3.4 |
| Total$_w$[b] (931) | 44.0 | 40.8 | 47.2 | 45.3 | 42.1 | 48.5 | 7.9 | 6.2 | 9.6 | 2.8 | 1.8 | 3.9 |

[a] One or more of the following: positive AIDT result, detectable anti-SARS-CoV-2 N IgG or detectable anti-SARS-CoV-2 RBG total antibodies in unvaccinated participants (in the absence of anti-N IgG, and history of a positive AIDT test.

[b] Total age/sex Valencia Community population weighted.

| Supplementary Table 4. Participants evaluated for the presence of SARS-CoV-2-Spike IFN- γ-producing T cells and Spike-binding neutralizing antibodies | | |
|---|---|---|
| **Parameter** | **Assessment of SARS-CoV-2-Spike T-cell immunity. no. of participants (%)** | **Assessment of neutralizing activity of sera against Wuhan Hu-1 and Omicron BA.1 and BA.2** |
| **Valencian Community Province** | | |
| Valencia | 76 (56) | 55 (55) |
| Alicante | 40 (29) | 40 (46) |
| Castellón | 21 (15) | 5 (5) |
| **Sex** | | |
| Male | 63 (46) | 50 (50) |
| Female | 74 (54) | 50 (50) |
| **Age group (range)** | | |
| 0-9 | 5 (3.6) | 5 (5) |
| 10-19 | 12 (8.8) | 8 (8) |
| 20-34 | 25 (18) | 19 (19) |
| 35-49 | 33 (24) | 24 (24) |
| 50-64 | 23 (17) | 16 (16) |
| 65-79 | 22 (16) | 14 (14) |
| >80 | 17 (12) | 14 (14) |
| **Vaccination/SARS-CoV-2 infection status** | | |
| Vaccinated/Experienced | 51 (37) | 34 (34) |
| Vaccinated/Naïve | 73 (53) | 55 (55) |
| Unvaccinated/Experienced | 11 (8) | 10 (10) |
| Unvaccinated/Naïve | 2 (1) | 1 (1) |
| Homologous booster dose | 36/88 (41) | 25/63 (40) |
| Heterologous booster dose | 52/88 (59) | 38/63 (60) |

| **Supplementary Table 5. Neutralizing antibodies and SARS-CoV-2-S-IFN-γ-producing T cells in vaccinated/SARS-CoV-2 experienced participants either boosted or not with an additional vaccine dose** | | | |
|---|---|---|---|
| Immunological parameter | Vaccinated/experienced with booster dose/ no. of participants evaluated | Vaccinated/experienced without booster dose/ no. of participants evaluated | *P* value |
| Neutralizing antibodies against Wuhan-Hu-1; median reciprocal $IC_{50}$ titer (range) | 2,610 (585-19,477)/12 | 1,446 (85-13,145)/22 | 0.10 |
| Neutralizing antibodies against Omicron BA.1; median reciprocal $IC_{50}$ titer (range) | 583 (19-36,637)/12 | 254 (10-16,974)/22 | 0.21 |
| Neutralizing antibodies against Omicron BA.2; median reciprocal $IC_{50}$ titer (range) | 42 (10-689)/12 | 149 (23-963)/22 | 0.56 |
| SARS-CoV-2-S-IFN-γ $CD8^+$ T cells; median frequency (range) | 0.52 (0-30)/21 | 0.19 (0-10)/30 | 1.0 |
| SARS-CoV-2-S-IFN-γ $CD4^+$ T cells; median frequency (range) | 0.08 (0-0.20)/21 | 0.07 (0-4)/30 | 0.32 |

**Supplementary Table 6.** Correlation of neutralizing antibody titers and SARS-CoV-2-S-IFN-γ-producing T-cell frequencies with the time elapsed since the last vaccine dose in vaccinated/SARS-CoV-2 experienced and vaccinated/SARS-COV-2 naïve participants.

| **Immunological parameter** | **Spearman rank test Rho value [range] (*P* value) in vaccinated/experienced** | **Spearman rank test Rho value [range] (*P* value) in vaccinated/naïve** |
|---|---|---|
| Neutralizing antibodies against Wuhan-Hu-1 | -0.26 [-0.55- 0.08] (0.13) | -0.34 [-0.55-0.07] (0.02) |
| Neutralizing antibodies against Omicron BA.1 | 0.06 [-0.29- 0.39] (0.74) | -0.39 [-0.59 -0.12] (0.004) |
| Neutralizing antibodies against Omicron BA.2 | -0.22 [-0.52- 0.13] (0.21) | -0.15 [-0.41, 0.12] (0.27) |
| SARS-CoV-2-S-IFN-γ $CD8^+$ T cells | -0.16 [-0.47- 0.19] (0.37) | -0.14 [-0.39-0.13] (0.31) |
| SARS-CoV-2-S-IFN-γ $CD4^+$ T cells) | -0.20 [-0.51- 0.15] (0.26) | 0.08 [-0.19-0.34] (0.57) |

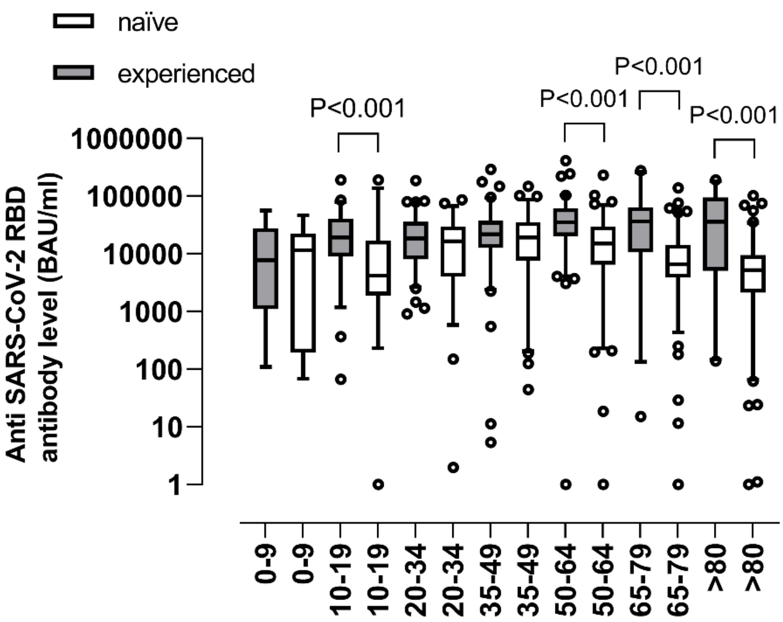
naïve
experienced
P<0.001
P<0.001
P<0.001
P<0.001
1000000
100000
10000
1000
100
10
1
Anti SARS-CoV-2 RBD antibody level (BAU/ml)
0-9
0-9
10-19
10-19
20-34
20-34
35-49
35-49
50-64
50-64
65-79
65-79
>80
>80
